# AI-based Detection of Central Retinal Artery Occlusion within 4.5 hours on Standard Fundus Photographs

**DOI:** 10.1101/2024.12.19.24319390

**Authors:** Ayse Gungor, Ilias Sarbout, Aubrey L. Gilbert, Steffen Hamann, Pierre Lebranchu, Cristina Hobeanu, Philippe Gohier, Catherine Vignal-Clermont, Oana M. Dumitrascu, Salomon-Yves Cohen, Wolf A. Lagrèze, Nicolas Feltgen, Frank van der Heide, Cédric Lamirel, Jost B. Jonas, Michael Obadia, Daniel Racoceanu, Dan Milea

## Abstract

**Background:** Prompt diagnosis of acute central retinal artery occlusion (CRAO) is crucial for therapeutic management and secondary prevention of associated neurological co-morbidities. However, most stroke centers lack on-site ophthalmic expertise prior to considering fibrinolytic treatments. We aimed to develop, train, and test a deep learning system (DLS) able to accurately detect hyper-acute CRAO on retinal color fundus photographs, during the critical treatment window of 4.5 hours after visual loss. We also evaluated the diagnostic performance of the DLS within 24 hours after visual loss, aiming to improve secondary prevention of stroke after CRAO.

**Methods:** Our retrospective, multicenter, multiethnic study included 1,322 color fundus photographs from 771 patients with various causes of acute visual loss, including CRAO, central retinal vein occlusion, non-arteritic anterior ischemic optic neuropathy, and healthy controls. Photographs were collected from 9 expert neuro-ophthalmology centers in 6 countries, including 3 randomized clinical trials. Training was performed on 1,039 photographs (517 patients), followed by testing on two datasets to discriminate CRAO cases at (i) hyper-acute stage (54 photographs, 54 patients) and (ii) within 24 hours after visual loss (110 photographs, 109 patients).

**Results:** The DLS achieved an area under the receiver operating characteristic curve (AUC) of 0.96 (95% confidence interval [CI], 0.95-0.98), a sensitivity of 92.6% (95% CI, 87.0-98.0), and a specificity of 85.0% (95% CI, 81.8-92.8) for detecting CRAO at hyper-acute stage, with similar results for CRAO diagnosis within 24 hours. The DLS outperformed neurologists on a subset of testing dataset at hyper-acute stage (120 photographs from 120 patients).

**Conclusions:** A DLS can accurately detect hyper-acute CRAO on retinal photographs within a time-window compatible with urgent fibrinolysis. Delayed diagnosis (24h) did not alter the ability of the DLS to accurately identify CRAO. If further validated, such systems could improve patient selection for fibrinolytic trials and optimize secondary stroke prevention.

**Clinical Trial Registration:** URL: https://www.clinicaltrials.gov; Unique identifier: NCT06390579.

## Introduction

Central retinal artery occlusion (CRAO) is considered an “eye stroke”,^1^ causing not only sudden visual loss, but also an increased risk for subsequent cerebrovascular and cardiovascular events, with a peak in the first 7 days.^2–4^ CRAO is a severe, blinding condition with fewer than 20% of patients experiencing meaningful visual recovery.^5,6^

Currently, no randomized clinical trial (RCT) has proven an effective treatment for CRAO,^7^ but early fibrinolytic intervention at hyper-acute stages (within 4.5 hours after visual loss) using intravenous (IV) alteplase has been associated with a recovery rate up to 50%, compared to 15.2% when treated between 4.5 and 6 hours after symptom onset.^8,9^ Preliminary results of the first RCT assessing IV fibrinolysis within 4.5 hours after visual loss have been recently released,^10^ indicating that there is currently a high need for additional RCTs to evaluate the efficacy of early fibrinolytic treatments in hyper-acute CRAO.^5^

CRAO diagnosis requires ophthalmic expertise, which is lacking in most stroke or emergency departments, making timely diagnosis and intervention difficult.^11^ Indeed, less than 40% of patients are initially evaluated by an ophthalmologist,^12–14^ possibly explaining why treatment is often initiated late in over 50% of cases,^15–17^ as more than half of the patients present after the critical 4.5-hour window.^18,19^ Non-ophthalmic physicians often lack confidence in diagnosing CRAO,^20^ knowing that fundus examination may appear normal in 5% of cases, further complicating diagnosis and treatment.^21^

The advent of artificial intelligence (AI) in medical imaging offers novel opportunities to develop tools for accurate and rapid diagnosis across a variety of medical conditions. Deep learning-based AI systems have already demonstrated excellent performance in the early recognition of stroke signs in prehospital settings,^22^ and the rapid identification of ST-elevation myocardial infarction.^23^ Recently, AI-based methods applied to retinal imaging has been used as a window to identify neurological disorders such as papilledema related to raised intracranial pressure,^24,25^ and dementia.^26^

Our study aimed to develop, train, and test a DLS for the early detection of CRAO on color fundus photographs: 1) within the critical 4.5-hour window to identify patients eligible for fibrinolysis, and 2) within the first 24 hours after visual loss, allowing improved secondary stroke prevention.

## Methods

### Ethics and Institutional Governance Approvals

This study was approved by ethical committees of each contributing center and the institutional review board (IRB00012801) of the coordinating center (Rothschild Foundation Hospital, Paris, France) and is registered on ClinicalTrials.gov (NCT06390579). It was conducted in accordance with the Declaration of Helsinki. This study followed the Standard Protocol Items: Recommendations for Interventional Trials–Artificial Intelligence (SPIRIT-AI)^27^, the Consolidated Standards of Reporting Trials–Artificial Intelligence (CONSORT-AI)^28^ and the Strengthening the Reporting of Observational Studies in Epidemiology (STROBE)^29^ reporting guidelines.

### Study Design

Color fundus photographs were collected from the clinical databases of 9 expert collaborating centers worldwide, including those of patients that were prospectively enrolled in CRAO RCTs, such as the European Assessment Group for Lysis in the Eye (EAGLE),^17^ Thrombolysis in Patients With Acute Central Retinal Artery Occlusion (THEIA),^10^ and TENecteplase in Central Retinal Artery Occlusion Study (TenCRAOS);^30^ as well as from two publicly available databases.^31,32^ These 11 databases included patients with diagnosis of sudden, painless visual loss (CRAO, central retinal vein occlusion [CRVO], non-arteritic anterior ischemic optic neuropathy [NAION]) and healthy controls. Inclusion criteria for patients with CRAO were specifically defined. In the training dataset, patients were eligible if the fundus photographs were taken within 30 days of symptom onset and whose diagnosis was confirmed by the referring expert. All patients with CRAO who were included in the testing datasets had the retinal imaging performed within 4.5 hours (and within 24 hours, respectively). Importantly, if CRAO photographs were taken after fibrinolysis, the patients were excluded. For all cases, including CRAO, CRVO, NAION, and healthy controls, photographs were excluded if they were taken in patients with unconfirmed diagnosis or in association with other retinal conditions or if they were of poor quality. Poor quality photographs were defined as those with artifacts, inadequate focus, poor illumination, or obscured retinal details that hindered accurate interpretation. As a result, out of the initially available 2,861 CRAO photographs and 827 non CRAO photographs, we excluded 2,538 CRAO photographs (88.7%) (Figure S1 in Supplement) and 253 non CRAO photographs (30.6%).

A DLS was developed, trained, and tested in a multiclass classification setting to differentiate CRAO from CRVO and NAION, as well as from photographs of healthy controls. The training and internal validation datasets consisted of 1,039 color fundus photographs: 614 photographs (517 patients) collected from 6 participating centers and 425 photographs from 2 publicly available databases. The participating centers contributed by providing 213 CRAO (164 patients), 125 CRVO (125 patients), 139 NAION (124 patients), and 137 normal (104 patients) photographs. We have also used photographs from public databases, with the following diagnosis: 16 CRAO, 66 CRVO, and 343 normal (Table 1). The first testing dataset, at hyper-acute stage, included 54 CRAO photographs (54 patients) taken within 4.5 hours after visual loss, 63 photographs (63 patients) with CRVO, 50 photographs (41 patients) with NAION, and 60 photographs (41 patients) of healthy controls. The second testing dataset included a total of 110 CRAO photographs (109 patients), whose photographs were captured within 24 hours after visual loss), 63 CRVO photographs (63 patients), 50 NAION photographs (41 patients), along with 60 photographs (41 patients) of healthy controls (Table 2). All photographs in the testing datasets were independent of those used in the training and internal validation datasets.

**Table 1.** Data used in the training and internal validation datasets

| Location | Center | No. of patients | No. of photographs used by classes | Age, mean (range), years | Sex, No. (%) |  |
| --- | --- | --- | --- | --- | --- | --- |
|  |  |  |  |  | Male | Female |
| Angers, France | Angers University Hospital | 157 | 80 normal<br>112 NAION | 71.6 (19-95) | 70 (44.6) | 73 (46.5) |
| Beijing, China | Beijing Institute of Ophthalmology | 6 | 6 normal<br>6 CRAO | 45.5 (30-59) | 4 (66.7) | 2 (33.3) |
| Freiburg, Germany | Eye Center, University of Freiburg | 30 | 50 CRAO | 62.2 (24-74) | 20 (66.7) | 10 (33.3) |
| Nagpur, India | Suraj Eye Institute | 19 | 19 normal<br>8 CRAO | 54.0 (33-71) | 5 (26.3) | 1 (5.3) |
| Paris, France | Ophthalmic Imaging and Laser Center | 68 | 48 CRVO<br>20 CRAO | 76.0 (46-97) | 33 (48.5) | 35 (51.5) |
|  | Rothschild Foundation Hospital | 237 | 32 normal<br>27 NAION<br>77 CRVO<br>129 CRAO | 65.2 (16-93) | 89 (37.5) | 68 (28.7) |
| Publicly Available Datasets | Rotterdam EyePACS<br>AIROGS | NA | 305 normal | NA | NA | NA |
|  | Kaggle | NA | 38 normal<br>66 CRVO<br>16 CRAO | NA | NA | NA |

Abbreviations: NAION, non-arteritic anterior ischemic optic neuropathy; CRAO, central
retinal artery occlusion; CRVO, central retinal vein occlusion; NA, not available.

**Table 2.**
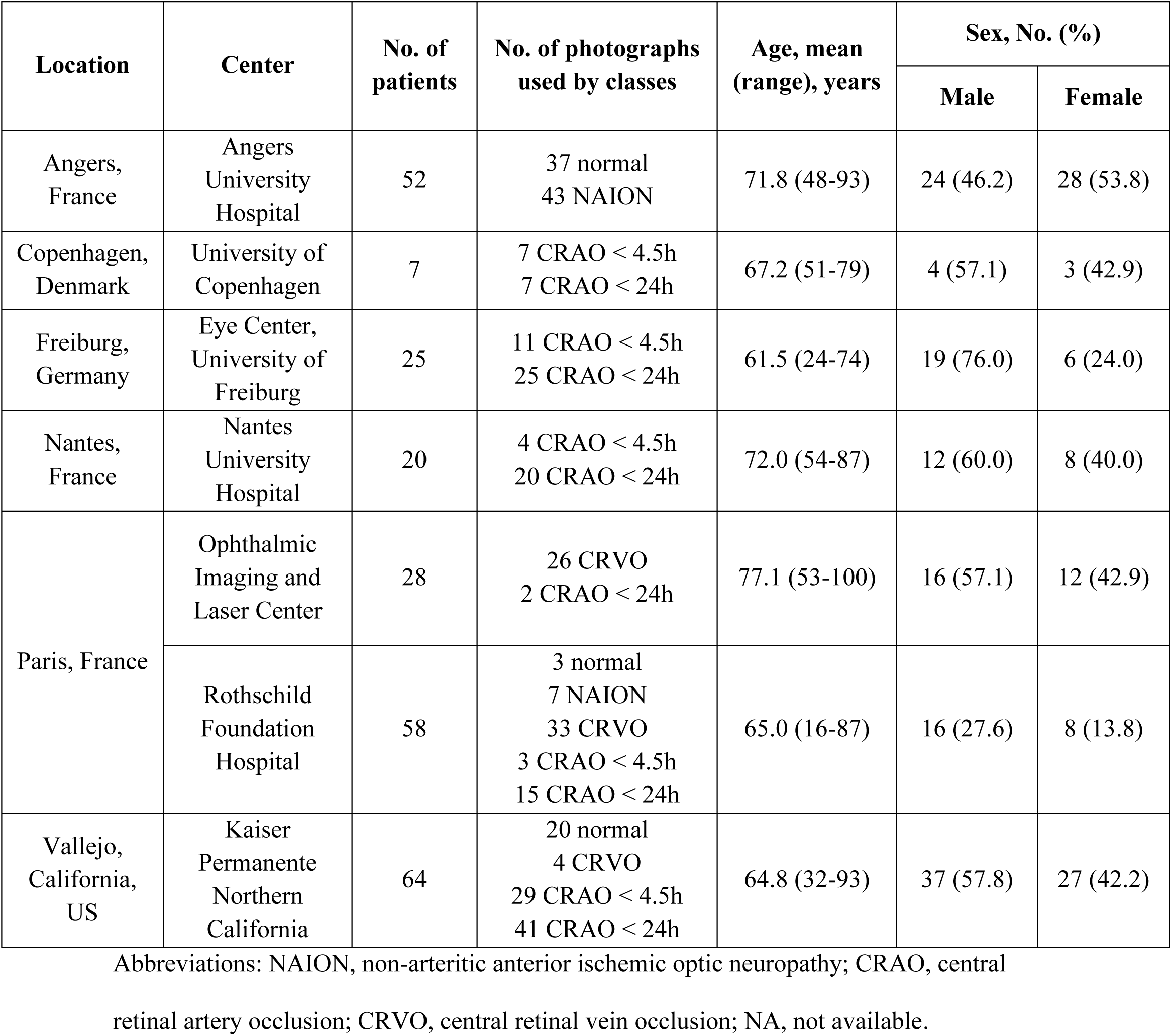
Data used in the testing datasets

| Location | Center | No. of patients | No. of photographs used by classes | Age, mean (range), years | Sex, No. (%) |  |
| --- | --- | --- | --- | --- | --- | --- |
|  |  |  |  |  | Male | Female |
| Angers, France | Angers University Hospital | 52 | 37 normal<br>43 NAION | 71.8 (48-93) | 24 (46.2) | 28 (53.8) |
| Copenhagen, Denmark | University of Copenhagen | 7 | 7 CRAO < 4.5h<br>7 CRAO < 24h | 67.2 (51-79) | 4 (57.1) | 3 (42.9) |
| Freiburg, Germany | Eye Center, University of Freiburg | 25 | 11 CRAO < 4.5h<br>25 CRAO < 24h | 61.5 (24-74) | 19 (76.0) | 6 (24.0) |
| Nantes, France | Nantes University Hospital | 20 | 4 CRAO < 4.5h<br>20 CRAO < 24h | 72.0 (54-87) | 12 (60.0) | 8 (40.0) |
| Paris, France | Ophthalmic Imaging and Laser Center | 28 | 26 CRVO<br>2 CRAO < 24h | 77.1 (53-100) | 16 (57.1) | 12 (42.9) |
|  | Rothschild Foundation Hospital | 58 | 3 normal<br>7 NAION<br>33 CRVO<br>3 CRAO < 4.5h<br>15 CRAO < 24h | 65.0 (16-87) | 16 (27.6) | 8 (13.8) |
| Vallejo, California, US | Kaiser Permanente Northern California | 64 | 20 normal<br>4 CRVO<br>29 CRAO < 4.5h<br>41 CRAO < 24h | 64.8 (32-93) | 37 (57.8) | 27 (42.2) |

Abbreviations: NAION, non-arteritic anterior ischemic optic neuropathy; CRAO, central
retinal artery occlusion; CRVO, central retinal vein occlusion; NA, not available.

### Development of the DLS

The DLS employed a convolutional neural network, optimized through a grid search on an internal validation set to determine the most effective combination of hyperparameters of the model, including batch size, learning rate, network size, cross-entropy weights, number of epochs and a set of data augmentation techniques. The architecture, detailed in Figure S2 in Supplement, included strided and classic convolutional layers, ReLU activation functions,^33^ fully connected layers and a SoftMax layer, which outputs a score for each of the four predicted classes.^34^ Dropout was used to prevent overfitting. To visualize the model’s decision-making process, the gradient-based approach Grad-CAM^35^ was employed and averaged to create disease-specific class-activation maps. These maps highlighted the areas with the highest pixel activation, helping to identify key image features that influenced the model’s decisions. This visual representation allowed us to align these regions with pathological cues typically assessed by experts. The performance of the DLS was calculated via the area under the receiver operating characteristic curve (AUC), sensitivity, specificity, accuracy and F-1 score.

### Preprocessing for Enhancing the DLS’s Robustness

Color fundus photographs are often captured under variable lighting conditions, resulting in uneven illumination caused by shadows, reflections, etc. affecting the quality of feature extraction. Such variations in colorimetric properties and contrast may also occur due to different camera models (Table S1 in Supplement). To address these challenges and enhance the robustness of our DLS, several preprocessing steps were implemented.

First, contrast-limited adaptive histogram equalization (CLAHE),^36^ a gold-standard preprocessing technique for fundus photographs, was applied to the luminance channel in the LAB color space to enhance local contrast. This method helped prevent over-amplification of noise in low-contrast areas by adaptively adjusting the contrast. Next, a novel approach for background removal was introduced, leveraging recent advancements in deep learning methodologies.^37^ Image dilation was applied followed by median blurring to estimate the background. This estimated background was then subtracted from the original image, effectively eliminating background color while preserving the integrity of the retinal structures (Figure 1). To further enhance the robustness of the DLS, various data augmentation techniques were randomly applied during the training, including random cropping, homographies, and adjustments to contrast and color.

**Figure 1.**
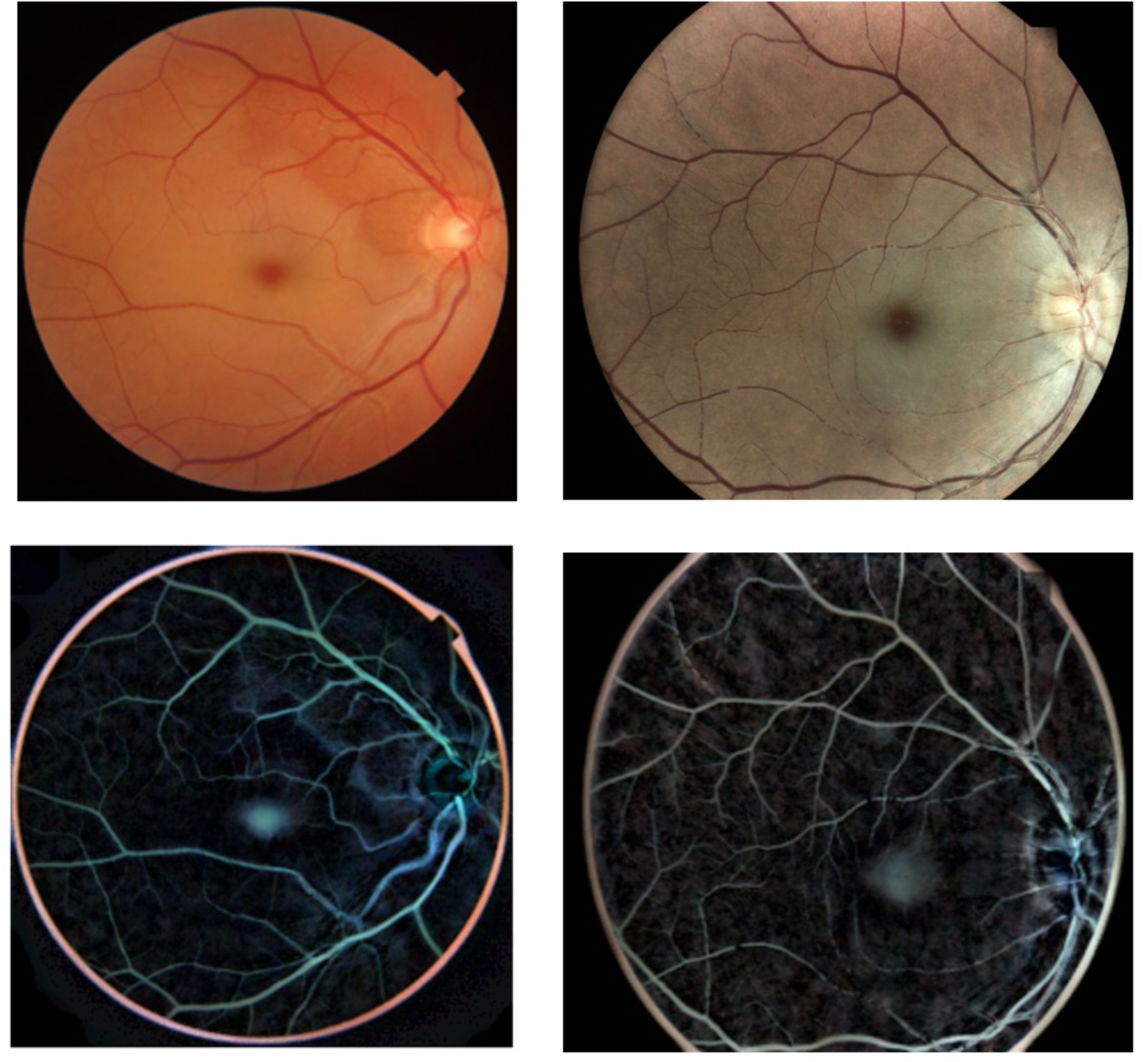
Preprocessing of the color fundus photographs. Original fundus photographs, taken with two different cameras in two patients with central retinal artery occlusion (top row). The bottom row represents the corresponding preprocessed images, after the custom background removal.

### Evaluation by Expert Neurologists for Comparative Analysis

We compared the performance of the DLS at 4.5 hours with that of three expert neurologists (G.A., C.S., and C.H.) Following a brief training session, each expert independently classified a subset of 120 color fundus photographs (120 patients) from the first testing dataset. This subset included 30 hyper-acute CRAO photographs (30 patients) taken within 4.5 hours after visual loss, 30 CRVO photographs (30 patients), 30 NAION photographs (30 patients), and 30 normal photographs (30 patients). The classifications were performed into four classes using the semi-automated image annotation application, Classif-Eye.^38^ The experts were masked to the patients’ clinical information and unaware of the classifications made by the DLS or the other experts. The overall agreement among the three experts was assessed using the Fleiss kappa score.^39^ The kappa agreement score was interpreted based on a previously published scale: 0 to 0.20 indicating no agreement, 0.21 to 0.39 minimal agreement, 0.40 to 0.59 weak agreement, 0.60 to 0.79 moderate agreement, 0.80 to 0.90 strong agreement, and scores exceeding 0.90 indicating almost perfect agreement.^40^ Additionally, p-values were calculated to compare the performance of the DLS and the experts, to highlight any significant differences in accuracy.

## Results

### Patient Characteristics

Demographic details were available for 630 of 771 included patients. The training and internal validation datasets consisted of 221 (42.7%) male individuals, 189 (36.6%) female individuals, with a mean (range) age of 68.1 (16-97) years (Table 1). The testing datasets included 128 (50.4%) male individuals and 92 (36.2%) female individuals, with a mean (range) age of 68.5 (16-100) years (Table 2). Demographic details by condition in the training and testing datasets can be found in Table S2 in the Supplement.

### Classification by the DLS and Neurologists

In a one-versus-rest model, the DLS distinguished CRAO cases from non-CRAO cases (including CRVO, NAION, and healthy controls) with an accuracy of 86.8% (95% CI, 77.4-92.6), misclassifying only 4 out of 54 CRAO photographs taken within 4.5 hours visual loss (Figure S3 in Supplement). The AUC for this task was 0.96 (95% CI, 0.95-0.98), with a sensitivity of 92.6% (95% CI, 87.0-98.0), a specificity of 85.0% (95% CI, 81.8-92.8), and a binary F-1 score of 76.9% (95% CI, 70.3-85.8). The performance at 24 hours after visual loss showed that the DLS achieved an AUC of 0.97 (95% CI, 0.96-0.99), a sensitivity of 94.5% (95% CI, 88.3-99.0), a specificity of 85.0% (95% CI, 81.8-92.8), an accuracy of 88.7% (95% CI, 78.5-95.9), and a binary F-1 score of 86.7% (95% CI, 83.4-93.7) (Figure S4 in Supplement). The averaged disease-specific class-activation maps highlighted the regions of interest with the highest pixel activation for each condition. In healthy controls, the entire retina showed diffuse pixel activation (Figure 2A). In CRAO cases, the highest pixel activation was in the macular region (Figure 2B). In CRVO cases, pixel activation was broadly distributed in the circumferential peripheral part of the macular region (Figure 2C), while in NAION cases, pixel activation was concentrated in the optic disc zone (Figure 2D).

**Figure 2.**
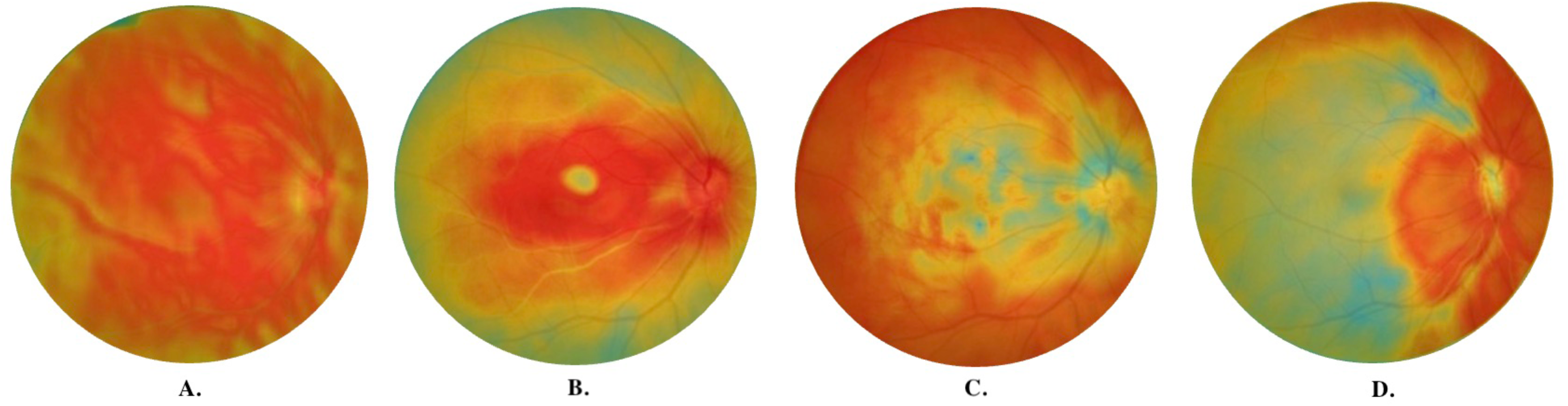
Superimposed and Averaged Disease-specific Class-activation Maps. Superimposed and averaged class-activation maps, revealing areas of interest with highest pixel activation to differentiate between **(A)** normal, **(B)** central retinal artery occlusion, **(C)** central retinal vein occlusion, and **(D)** non-arteritic anterior ischemic optic neuropathy.

Among the 199 patients included at less than 4.5 hours after visual loss, a subset of 120 patients (30 CRAO, 30 CRVO, 30 NAION and 30 healthy controls) were evaluated by both the DLS and the experts. The DLS detected all 30 CRAO cases, achieving an accuracy of 85% (95% CI, 71.3-90.8), sensitivity of 100% (95% CI, 88.4-100.0), and specificity of 80% (95% CI, 70.1-85.3). In comparison, the experts’ performance on the same task showed accuracy ranging from 66.7% to 79.2%, sensitivity from 50% to 63.3%, and specificity from 72.2% to 84.4% (Figure 3). The classifications made by both the DLS and the experts, including accurate classifications and misclassifications for CRAO and non CRAO cases, are provided in Figure S5 in Supplement. The intergrader agreement was moderate, with a kappa score of 0.73. Disagreement among neurologists was noted in 18 photographs, including 5 (27.7%) photographs of hyper-acute CRAO. Statistical analysis using McNemar test^41^ disclosed statistically significant differences between the DLS and experts 2 and 3 (p-values < 0.05), but no significant difference (p-value > 0.05) between the DLS and the expert 1. These results indicate that the DLS performed similarly or better than the experts in detecting CRAO at the hyper-acute stage.

**Figure 3.**
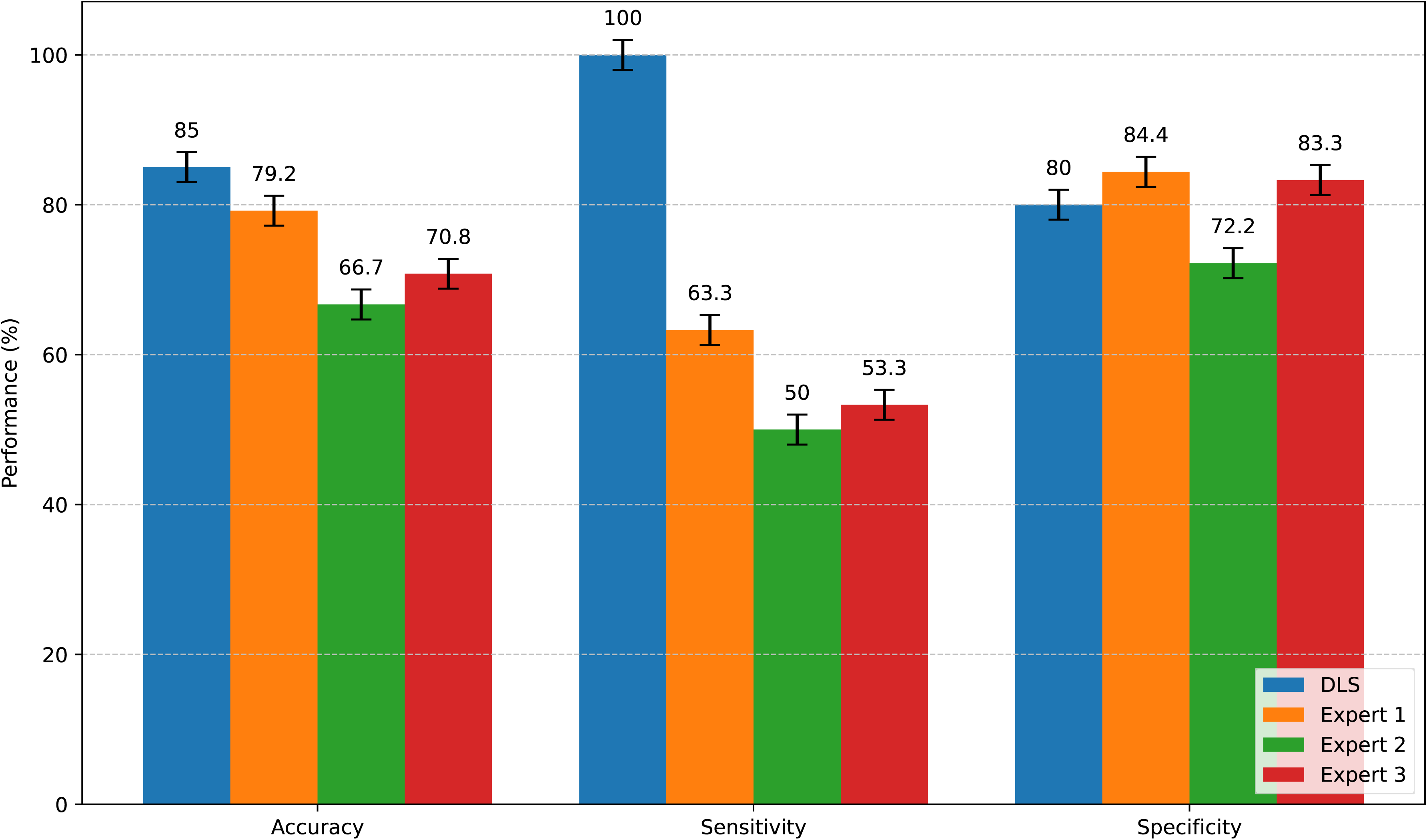
Performance (Accuracy, Sensitivity, Specificity) of the Deep Learning System and the 3 Neurologists in Detecting Hyper-acute Central Retinal Artery Occlusion. Comparison of the performance (accuracy, sensitivity, specificity) of the deep learning system to the performance of each expert in detecting hyper-acute central retinal artery occlusion (CRAO) from other differential diagnosis of sudden, painless visual loss (central retinal vein occlusion and non-arteritic anterior ischemic optic neuropathy) and from healthy controls on 120 color fundus photographs (30 hyper-acute CRAO and 90 non CRAO cases).

## Discussion

The main finding of our study is that a DLS can effectively identify CRAO at both hyper-acute (4.5 hours) and delayed (24 hours) stages, distinguishing it from photographs of healthy controls and from other differential diagnosis of sudden, painless visual loss such as CRVO and NAION. The DLS accurately identified CRAO cases, missing only 4 out of 54 CRAO photographs (7.4%) taken within 4.5 hours and 6 out of 110 photographs (5.4%) taken within 24 hours after visual loss. The performance of the DLS was superior to that of neurologists, suggesting that such a system could potentially serve an assistive aid in an emergency setting.

Superimposed and averaged disease-specific class-activation maps highlighted the regions of interest with the highest pixel activations, indicating the areas of the images most critical for the DLS’s predictions. This method has already been used as an innovative way to determine areas of interest in similar diagnostic applications.^42^ In healthy controls, the DLS established the diagnosis using averaged class-activation maps which were distributed over the whole retina (Figure 2A). Conversely, in CRAO cases the highest area of interest was limited to the macular region (Figure 2B), which is indeed preferentially affected in CRAO. In CRVO cases, pixel activation was broadly distributed in the circumferential peripheral part of the macular region, reflecting the widespread retinal hemorrhages (Figure 2C). Lastly, in NAION, the region of interest was unsurprisingly concentrated in the region of the optic nerve head, where hemorrhages and edema are clinically seen (Figure 2D). Thus, the areas highlighted by the DLS’s superimposed and averaged class-activation maps align with the primary anatomical sites affected by each specific condition.

The overall accuracy of the DLS was 87% in detecting CRAO at the hyper-acute stage. Importantly, many of the CRAO photographs in this testing dataset were collected prospectively in validated prospective RCTs, evaluating fibrinolytic treatments in this condition.^10,17,30^ Identification of patients at the hyper-acute stage remains challenging, primarily due to delayed presentation and/or CRAO diagnosis, which is one of the reasons for delayed inclusions. Indeed, in the EAGLE trial, the mean time from symptom onset to presentation was 9.5 hours, with a mean of 11 hours to treatment.^17^ Our study, which represents one of the largest CRAO studies to day, suggests that such an automated assistive system may improve early identification of CRAO in the future, with therapeutic implications.

### Limitations

Our study has inherent limitations. The training was performed on a retrospectively collected dataset study, excluding those with multiple ophthalmic pathologies, limiting the study’s ability to generalize its findings to real life situations. We used standard fundus photography as a method to identify CRAO. While optical coherence tomography is a more sensitive modality for detecting acute CRAO, capable of identifying subtle retinal changes not visible on fundus photographs,^21^ it was not included in this study due to its high cost and the expertise required for image acquisition and interpretation, which limit its availability in many emergency settings. Additionally, we only used photographs taken with traditional cameras, which means that data acquired with wide-angle and handheld cameras will need further investigation.

## Conclusion

A DLS can detect CRAO at both the hyper-acute (within 4.5 hours after visual loss) and the delayed stage (within 24 hours) using color fundus photographs. Beyond aiding in CRAO diagnosis, the DLS can also improve patient selection for fibrinolytic trials and secondary stroke prevention. Future prospective studies are necessary to validate the performance of the DLS, at best in real-life conditions.

## Data Availability

Data can be shared on reasonable requests.

## Acknowledgements

We thank VISIO Foundation, France for supporting this study; as well as Bouchra Touzani, MSc and Emmanuel Blondel, MSc at the Rothschild Foundation Hospital for their valuable assistance during this study.

